# Combining A Massively Parallel Reporter Assay and Human Data to Elucidate Genetic Mechanisms Driving Risk for Juvenile Idiopathic Arthritis

**DOI:** 10.64898/2026.08.24.26361225

**Authors:** Kaiyu Jiang, Susan Kales, Ryan Tewhey, James N. Jarvis

## Abstract

**Background:** While progress has been made in identifying the true risk-driving single nucleotide polymorphisms (SNPS) on juvenile idiopathic arthritis (JIA) risk haplotypes, the affected cells and target genes largely remain unknown.

**Methods:** We used data from a previously published massively parallel reporter assay (MPRA) to query human data in the Database of Immune Cell eQTLs (DICE) and the Gene-Tissue Expression (GTEx) database to identify affected cells and target genes of MPRA-identified SNPs in immune cells and relevant tissues.

**Results:** SNPs identified on MPRA were associated with gene expression levels in a broad range of immune cells in the DICE database, including CD4+ and CD8+ T lymphocytes, monocytes, NK cells, and B cells. MPRA-identified SNPs showed strong associations with gene expression in GTEx whole blood, spleen, and/or EBV-transformed lymphocytes.

**Conclusions:** Our data show the efficacy of combining MPRA and using human cells/tissue expression data to elucidate complex mechanisms driving genetic risk for JIA.

## 1. Introduction

Juvenile idiopathic arthritis (JIA) is one of the most common chronic illnesses affecting children in North America (2). The pathogenesis of JIA is poorly understood, but clinical and experimental evidence supports the idea that JIA is driven by complex, and probably aberrant, interactions between the innate and adaptive immune systems(3) (4).

Genetic variants are well-established drivers of risk for JIA. In a study of the Utah population database, for example, Prahalad and colleagues showed that the relative risk for JIA in first degree relatives of affected patients is nearly12 times the risk in the general population, and that for second degree relative relatives >5 times (5). Genome wide scans have identified > 25 regions (outside of the HLA locus) that confer risk for JIA (6–8). However, translating these findings to an understanding of pathobiology and/or to inform care has been challenging. Three impediments have slowed such efforts: (1) the SNPs identified on genome-wide scans are each in linkage disequilibrium (LD) with dozens or hundreds of other SNPs, complicating the identification of the true risk-driving variants; (2) most of the LD blocks encompassing the JIA risk loci encompass multiple genes and non-coding elements, complicating the identification of the affected genes at each locus; (3) the 3D structure of the genome further complicates the identification of target genes. Pelikan and colleagues have shown, for example, that many target genes for systemic lupus risk variants aren’t actually on the risk haplotypes(9). Further complicating efforts to translate GWAS results to elucidate biology and/or inform clinical medicine is the fact that, for most autoimmune diseases, genetic risk has been shown to largely affect non-coding, regulatory functions (4, 10). Thus, the single nucleotide polymorphisms (SNPs) that drive risk are more likely to impact levels of expression of pathologically relevant genes than to alter the coding sequences (and functions) of such genes (although such instances have been well documented(11).

We have attempted to disentangle LD to identify the true risk-driving SNPs on the JIA risk haplotypes by using <u>massively</u> parallel reporter assays (MPRA)(12). These assays use oligonucleotide reporters to assess the *intrinsic* capacity of one allele or another of the tested SNPs to alter expression of the green fluorescence protein reporter. Thus, MPRA measure a physiologically-relevant feature of the tested genetic variants. Furthermore, MPRA can disentangle LD because they test intrinsic properties of each allele, independent of LD. However, MPRA do not test SNPs/alleles in a true genomic context, and, thus, may generate both false positives and false negatives. Thus, while MPRA are an excellent screening procedure to sift through the tens of thousands of genetic variants on autoimmune risk haplotypes (there are 14,758 SNPs on the known, non-HLA JIA risk haplotypes) to prioritize them for further laboratory characterization, the relevance of the identified SNPs to the disease being queried requires further investigation. In this study, we show that existing human data can be used to further elucidate MPRA findings and add insight into <u>candidate</u> target genes, affected cells and tissues, and immunopathology.

## 2. Materials and Methods

### 2.1. Massively parallel reporter assay (MPRA) in K562 cells

We performed MPRA in K562 cells and K562 cells stimulated with interferon-gamma (IFNγ). Details of these assays have been published(12) and are summarized briefly here. We queried SNPs within LD blocks defined by Hinks(6) and Hersh(13), using a cut-off of r^2^ = 0.80 to choose those SNPs in strong LD with the index SNPs. The regions queried are shown in **Table 1**. We tested 7,312 b(1)i-allelic SNPs. We then prepared and characterized oligonucleotide libraries was following published methods(14). Oligos were synthesized as 230 bp sequences containing 200 bp of genomic sequence and 15 bp of adapter sequence at either end (5’ACTGGCCGCTTGACG [200 bp oligo] CACTGCGGCTCCTGC3’). Unique 20 bp barcodes were added by PCR along with additional constant sequence for incorporation into a backbone vector by gibson assembly. The oligo library was expanded in E.coli and sequenced using Illumina 2×150 bp chemistry to acquire barcode/oligo pairings. The library underwent restriction digest and green fluorescence protein (GFP) with a minimal TATA promoter inserted by gibson assembly resulting in the 230 bp oligo sequence positioned directly upstream of the promoter and the 20 bp barcode falling in the 3′ UTR of GFP. After expansion within E.coli the final plasmid library was sequenced by Illumina 1×30 bp chemistry to acquire a baseline representation of each oligo within the library. The oligonucleotide libraries were electroporated into K562 cells, with or without IFNγ (250 ng/ml), a pathologically relevant dose for JIA pathobiology (12, 15). Green fluorescence protein RNA, tagged with bar codes for each allele, was isolated and sequenced as described in(12).

**Table 1:** Regions Queried in the K562 MPRA.

| **Locus Name** | **Tag SNP** | **GRCh38/hg38 Haplotype** |
| --- | --- | --- |
| PTPN22 | rs6679677 | chr1:113,761,186-113,834,946 |
| STAT4 | rs10174238 | chr2:191,079,016-191,108,308 |
| PTPN2 | rs2847293 | chr18:12,774,327-12,809,341 |
| ANKRD55 | rs71624119 | chr5:56,141,024-56,146,422 |
| IL2-IL21 | rs1479924 | chr4:122,151,854-122,619,603 |
| TYK2 | rs34536443 | chr19:10,317,045-10,381,598 |
| IL2RA | rs7909519 | chr10:6,028,313-6,055,320 |
| SH2B3-ATXN2 | rs3184504 | chr12:111,395,984-111,645,358 |
| ERAP2-LNPEP | rs27290 | chr5:96,884,383-97,038,046 |
| UBE2L3 | rs2266959 | chr22:21,556,931-21,628,971 |
| C5orf56/IRF1 | rs4705862 | chr5:132,477,527-132,496,822 |
| RUNX1 | rs9979383 | chr21:35,323,611-35,365,944 |
| IL2RB | rs2284033 | chr22:37,135,077-37,141,474 |
| ATP8B2/IL6R | rs11265608 | chr1:154,319,242-154,406,893 |
| ZFP36L1 | rs12434551 | chr14:68,784,174-68-794,755 |
| JAK1 | rs10889504 | chr1:64,924,820-64,975,081 |
| PRR9/LOR | rs873234 | chr1:153,249,128-153,270,022 |
| PTH1R | rs1138518 | chr3:46,889,988-46,932,682 |
| ILDR1/CD86 | rs111700762 | chr3:122,023,892-122,102,059 |
| AHI1/LINC00271 | rs9321502 | chr6:135,303,673-35,371,709 |
| HBP1 | rs111865019 | chr7:107,156,092-107,390,877 |
| WDFY4 | rs1904603 | chr10:48,776,493-48,805,795 |
| RNF215 | rs57533109 | chr22:30,289,571-30,403,731 |

For each replicates, we extracted the first 20bps from the sequenced reads and used them to assign reads to either reference or alternative allele for variants based on our barcode library. We then generated a count-table for each variant, including the names of variants, as well as the counts of the reference and alternative alleles for each replicate. After merging replicates, we generated a master count-table for each allele for each of the replicate of DNA (plasmid library), RNA expression in K562 cells, and RNA expression in K562 cells with IFNγ treatment. We used a customized R script based on that described by Tewhey(14) to process the count-table and identify the DNA elements, defined as a region on chromosome that is investigated in MPRA, with active regulatory activities and the variants that can alter the regulatory activities. We used DESeq2(16), to normalize counts and fit the dispersion using ‘local fit’. The distribution of the log2 fold changes between RNA and DNA were investigated. Since we found the distribution was not centered at 0, we adjusted the size factors for DNA and RNA samples according to the offset, then repeated the normalization. Oligos showing differential RNA expression relative to the plasmid DNA were then identified by the nbinomWaldTest function in DESeq2 (a Wald Test for the coefficients in a negative binomial generalized linear model) and applying a threshold of 0.01 for the False Discovery Rate and a minimum fold change of 1.5. For DNA elements that displayed significant regulatory activity, we applied a t-test on the log2-transformed RNA/plasmid ratios for each paired replicate (e.g. alternative alleles of K562 replicate 1 vs reference alleles of K562 replicate 1) to determine whether the reference and alternate allele had a similar activity. The p-values from t-test were adjusted through the Benjamini-Hochberg process into FDR values, applying a cutoff of FDR 0.01 to call those variants showing altered regulatory activities between alleles.

Raw data from the MPRA sequencing has been deposited in the National Library of Medicine’s Sequence Read Archive (SRA), BioProject accession number PRJNA818294.

### 2.2 Querying the Database of Immune Cell Expression Quantitative Trait Loci (DICE)

The Database of Immune Cell eQTLs is a publicly-available database comprising RNAseq data from 13 different immune cell types from genotyped donors (17, 18). The database was last updated in February, 2022 and contains data from 91 individuals. For each SNP identified by MPRA, we queried DICE data to identify eQTLs associated with each SNP; discovery of eQTLs in the original DICE project was undertaken using the Matrix eQTL package(19) and is described in detail in(18). We considered adjusted p-values of 0.05 as reflecting a significant associated between the expression of a specific gene within a given cell type and the queried SNP.

### 2.2. Querying the Gene-Tissue Expression (GTEx) Database

The Gene-Tissue Expression project database (V10 [dbGaP Accession phs000424.v10.p2]-https://gtexportal.org/home/tissueSummaryPage) contains tissue-level gene expression data from 945 genotyped, postmortem donors(20). The GTEx eQTL calculator allows users to query the GTEx data to identify eQTLs within any of the GTEx cells/tissues using SNP (RSID) and ENSEMBL gene name as input. Using the SNPs identified on MPRA, we queried three GTEx tissue comprised of or enriched for immune cells: whole blood, EBV-transformed lymphocytes, and spleen. We also queried expression levels in liver because of the liver’s importance in synthesizing a broad range of inflammatory mediators. We identified the candidate target genes using either the DICE data (where eQTLs were identified) or using publicly available 3D chromatin data(21). GTEx eQTL data are adjusted for multiple comparisons and provide a threshold for significance for each cell type or tissue.

## 3. Results

As reported previously(12), we identified n=42 unique SNPs in MPRA assays performed in unstimulated K562 cells, and another n=42 unique SNPs in K562 cells stimulated with IFNγ. These SNPs are listed in **Tables 2** and **3**.

**Table 2.**
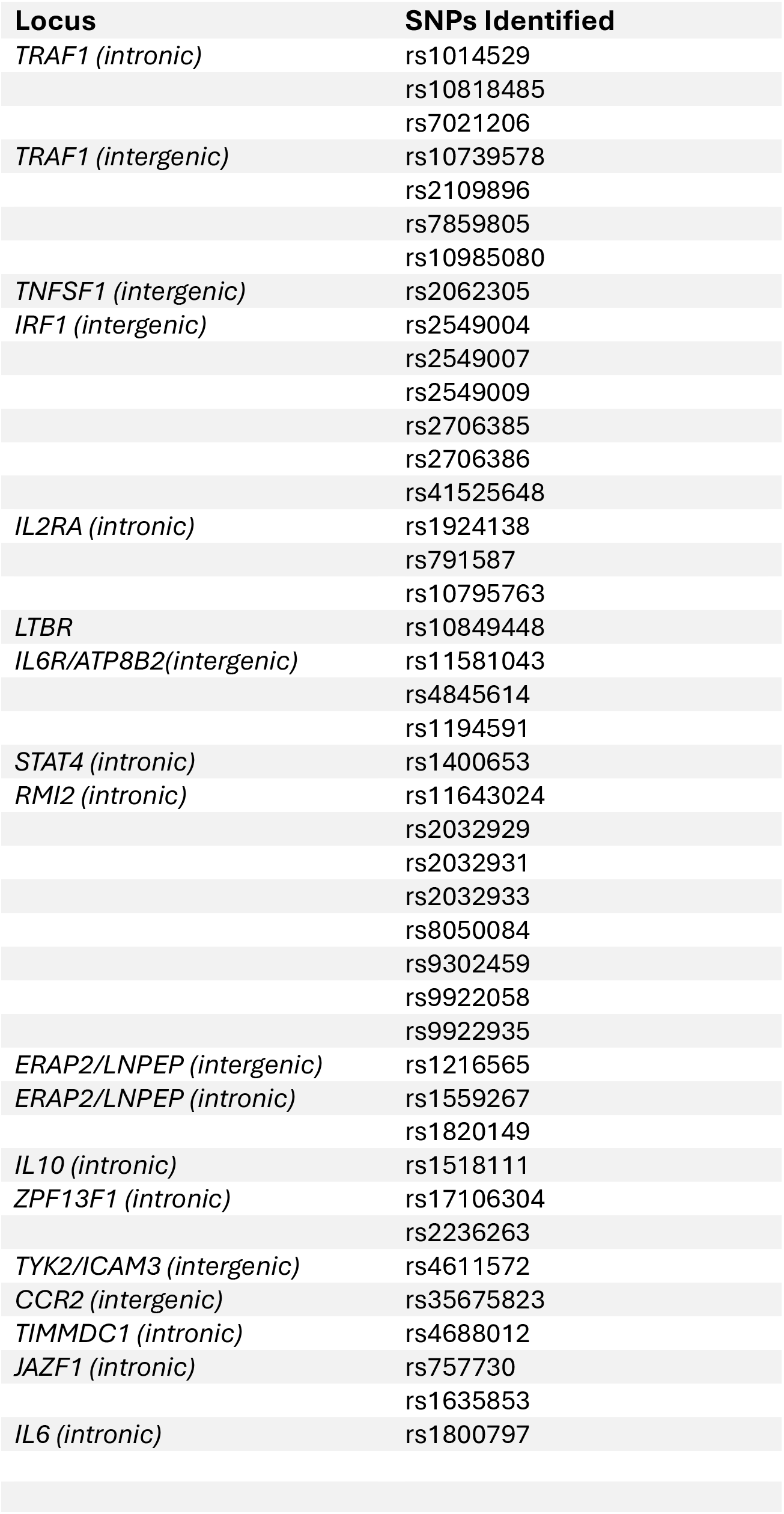
Unique SNPs Screening Positive in Unstimulated K562 Cells (n=42)

**Table 3.** Unique SNPs Screening Positive in K562 Stimulated By Interferon Gamma (n=42)

| **LOCUS** | **SNP(s)** |
| --- | --- |
| *PTPN2 (intronic)* | rs1217378 |
| *IL6/ATP8B2 (intronic)* | rs11581043 |
|  | rs1194608 |
| *IL6/ATP8B2 (intergenic)* | rs4845369 |
| *IL10 (intronic)* | rs1518111 |
|  | rs2945417 |
| *STAT4 (intronic)* | rs1996400 |
| *CCR2 (intergenic)* | rs7374671 |
|  | rs4683215 |
|  | rs35053103 |
|  | rs2888523 |
|  | rs2888524 |
|  | rs34997146 |
|  | rs62242985 |
|  | rs6441972 |
| *TIMMDC1 (intronic)* | rs4687853 |
|  | rs2177812 |
|  | rs7610049 |
| *ERAP2/LNPEP (intergenic)* | rs193994 |
|  | rs1216565 |
| *ERAP2/LNPEP (intronic)* | rs430827 |
| *IRF1 (intergenic)* | rs41525648 |
|  | rs2706385 |
|  | rs2706386 |
| *TRAF1 (intergenic)* | rs10818481 |
|  | rs2900180 |
| *IL2RA* | rs791592 |
| *ATXN2 (promoter)* | rs10849962 |
| *ATXN2 (exonic enhancer?)* | rs6416335 |
| *ZFP36L1 (intronic)* | rs3742887 |
| *ZFPL1 (intergenic)* | rs1595260 |
|  | rs4899258 |
| *RMI2 (intronic)* | rs34764020 |
|  | rs7205578 |
|  | rs6498184 |
| *PTPN2 (intergenic)* | rs4327116 |
|  | rs2847279 |
|  | rs9952753 |
| *PTPN2 (intronic)* | rs515151 |
| *TYK2/ICAM3 (intronic)* | rs2278442 |
|  | rs2304240 |
| *TYK2/ICAM3 (intergenic)* | rs510506 |

### 3.1. Analyses from the DICE Database

To determine whether any of these SNPs might influence gene expression in human immune cells, we queried the DICE data base. Of the n=42 SNPs identified by MPRA in unstimulated K562 cells, n=12 (28%) were also identified as eQTLs within the DICE database. These SNPs, and the cells in which they are associated with transcriptional effects, are shown in **Table 4**. The table also provides allelic information with transcription levels for each cell type showing significant association. Similarly, of the n=42 SNPs identified in IFNγ-stimulated K562 cells, n=26 (62%) were identified as eQTLs within the DICE database. These SNPs, and the cells in which they are associated with transcriptional effects, are shown in **Table 4**. Overall, 38% of the SNPs identified on the MPRAs as having intrinsic effects on gene expression were also shown to be associated with expression levels of relevant candidate target genes in the DICE database.

**TABLE 4:** DICE eQTLs for MPRA-Screened SNPs: Unstimulated K562 Cells. Abbreviations: TFH – T follicular helper cells TREG – Regulatory T cells NK – Natural Killer

| SNP | Alleles | Gene | Cells Impacted (adjusted p=values) - Allelic Effects |
| --- | --- | --- | --- |
| rs1014529 | Reference allele: C<br>Alternate allele: G | *TRAF1* | **C>G**<br>CD4+ TFH (p=0.0017)<br>CD4+ naïve TREG (p=0.027)<br>CD4+ memory TREG (p=0.032) |
| “ | “ | *PSMD5-AS1*<br>(ENSG00000226752) | **C>G**<br>CD4+ naïve-activated (p=0.0020)<br>CD4+ naïve (p=0.0025)<br>CD4+ Th2 (p=0.0049)<br>CD4+ memory TREG (p=0.0065)<br>CD8+ naïve (p=0.0092)<br>Monocyte, M2 (p=0.015)<br>CD4+ Th1/Th17 (p=0.016)<br>CD4+ Th17 (p=0.016)<br>CD8+ naïve, activated (p=0.017)<br>CD4+ naïve TREG (p=0.017)<br>B cell, naïve (p=0.026)<br>CD4+ TFH (p=0.039) |
| rs10818485 | Reference allele: G<br>Alternate allele: C | *TRAF1* | **G>C**<br>CD4+ TFH (p=0.017)<br>CD4+ TREG naïve (p=0.028)<br>CD4+ memory TREG (p=0.032) |
| “ | “ | *PSMD5-AS1*<br>(ENSG00000226752) | **G>C**<br>CD4+ naïve-activated (p=0.0020)<br>CD4+ naïve (p=0.0025)<br>CD4+ Th2 (p=0.0049)<br>CD4+ memory TREG (p=0.0065)<br>CD8+ naïve (p=0.0092)<br>Monocyte, M2 (p=0.015)<br>CD4+ Th1/Th17 (p=0.016)<br>CD4+ Th17 (p=0.016)<br>CD8+ naïve, activated (p=0.017)<br>CD8+ naïve, activated (p=0.017)<br>CD4+ naïve TREG (p=0.017)<br>B cell, naïve (p=0.026)<br>CD4+ TFH (p=0.039) |
| rs7021206 | Reference allele: G<br>Alternate allele: A | *TRAF1* | **G>C**<br>CD4+ TFH (p=0.017)<br>CD4+ TREG naïve (p=0.028)<br>CD4+ memory TREG (p=0.032) |
| “ | “ | *PSMD5-AS1*<br>(ENSG00000226752) | **G>A except where noted**<br>CD4+ naïve-activated (p=0.0020)<br>CD4+ naïve (p=0.0025)<br>CD4+ Th2 (p=0.0049)<br>CD4+ memory TREG (p=0.0065)<br>CD8+ naïve (p=0.0092)<br>Monocyte, M2 (p=0.015) **A>G**<br>CD4+ Th1/Th17 (p=0.016)<br>CD4+ Th17 (p=0.016)<br>CD8+ naïve, activated (p=0.017)<br>CD8+ naïve, activated (p=0.017) **A>G**<br>CD4+ naïve TREG (p=0.017) |
|  |  |  | B cell, naïve (p=0.026)<br>CD4+ TFH (p=0.039) |
| **SNP** |  | **Gene** | **Cells Impacted (adjusted p=values) – Allelic effects** |
| rs10739578 | Reference allele: G<br>Alternate allele: T | *TRAF1* | **C>T**<br>CD4+ TFH (p=2.0e-9)<br>CD4+ Th2 (p=0.0000061)<br>CD4+ naïve TREG (p=0.000015)<br>CD4+ naïve (p=0.000017)<br>CD4+ memory TREG (p=0.000020)<br>CD4+ Th17. (p=0.000030)<br>CD4+ Th1 (p=0.0010)<br>CD4+Th1/Th17 (p=0.0021)<br>CD8+ naïve (p=0.0091)<br>B cell naïve (p=0.0030) |
| rs10739578 (cont'd) |  | *PSMD5-AS1*<br>(ENSG00000226752) | **C>T**<br>CD4+ naïve, activated (p=0.000016)<br>CD4+ naïve (p=0.000043)<br>CD4+ memory TREG (p=0.000048)<br>CD8+ naïve (p=0.000087)<br>CD4+ Th2 (p=0.00010)<br>B cell naïve (p=0.00010)<br>CD4+ Th17 (p=0.00016)<br>CD4+ naïve TREG (p=0.00018)<br>CD4+ TFH (p=0.00034)<br>CD4+ Th1/Th17 (p=0.00036)<br>Monocyte, M2 (p=0.00040)<br>CD8+ naïve, activated (p=0.00078)<br>CD4+ Th1 (p=0.0011)<br>Monocyte, classical (p=0.0013)<br>NK CD56dimCD16+ (p=0.011) |
| “ |  | *C5* | **C>T**<br>B cell naïve (p=0.0026)<br>CD4+ naïve (p=0.0046)<br>Monocyte, M2 (p=0.0066)<br>Monocyte, classical (p=0.033)<br>CD8+ naïve (p=0.034) |
| rs2109896 | Reference allele: C<br>Alternate allele: T | *TRAF1* | **C>T**<br>CD4+ TFH (p=0.0017)<br>CD4+ naïve TREG (p=0.028)<br>CD4+ memory TREG (p=0.032) |
| “ |  | *PSMD5-AS1*<br>(ENSG00000226752) | **C>T**<br>CD4+ naïve, activated (p=0.0020)<br>CD4+ naïve (p=0.0025)<br>CD4+ Th2 (p=0.0049)<br>CD4+ memory TREG (p=0.0065)<br>CD8+ naïve (p=0.0092)<br>Monocyte, M2 (p=0.015)<br>CD4+ Th1/Th17 (p=0.0016)<br>CD4+ Th17 (p=0.016)<br>CD8+ naïve, activated (p=0.016)<br>CD4+ naïve TREG (p=0.017)<br>B cell naïve (p=0.026)<br>CD4+ TFH (p=0.039) |
| rs7859805 | Reference allele: A<br>Alternate allele: G | *TRAF1* | **A>G**<br>CD4+ TFH (p=0.0017)<br>CD4+ naïve TREG (p=0.028)<br>CD4+ memory TREG (p=0.032) |
| “ |  | *PSMD5-AS1*<br>(ENSG00000226752) | **A>G**<br>CD4+ naïve, activated (p=0.0020)<br>CD4+ naïve (p=0.0025)<br>CD4+ Th2 (p=0.0049) |

|  |  |  | CD4+ memory TREG (p=0.0065)<br>CD8+ naïve (p=0.0092)<br>Monocyte, M2 (p=0.015)<br>CD4+ Th1/Th17 (p=0.0016)<br>CD4+ Th17 (p=0.016)<br>CD8+ naïve, activated (p=0.016)<br>CD4+ naïve TREG (p=0.017)<br>B cell naïve (p=0.026)<br>CD4+ TFH (p=0.039) |
| --- | --- | --- | --- |
| SNP |  | Gene | Cells Impacted (adjusted p=values) – Allelic effects |
| rs11643024 | Reference allele: A<br>Alternate allele: G | *RP11-485G7.6*<br>(ENSG00000262703) | B cell naïve (p=0.034) **A>G** |
| “ |  | *RP11-485G7.5*<br>(ENSG00000263080) | B cell naïve (p=0.0011) **A>G** |
| rs1216565 | Reference allele: C<br>Alternate allele: A | *ERAP2* | **A>C**<br>CD4+ naïve TREG (p=1.6e-24)<br>CD4+ TFH (p=6.6e-24)<br>NKcell CD56dimCD16+ (p=1.5e-23)<br>CD4+ Th2 (p=4.4e-23)<br>Monocytes, classical. (p=9.6e-22)<br>CD4+ memory TREG (p=1.3e-21)<br>CD4+ Th17 (p=2.6e-21)<br>CD4+ Th1/Th17 (p=3.0e-21)<br>Monocyte, M2 (p=9.2e-21)<br>CD8+ naïve (p=8.7e-19)<br>CD4+ naïve (p=1.1e-18)<br>CD4+ Th1 (p=9.9e-18)<br>CD8+ naïve (activated) (p=1.5e-13)<br>B cell naïve (p=1.1e-12)<br>CD4+ naïve (activated) (p=9.0e-12) |
| rs1216565 (cont'd) |  | *CTD-2260A17.1*<br>(ENSG00000248734) | CD4+ naïve (p=0.00070) **A>C** |
| rs1559267 | Reference allele: A<br>Alternate allele: T | *ERAP2* | **T>A**<br>CD4+ naïve TREG (p=1.6e-24)<br>CD4+ TFH (p=6.6e-24)<br>NKcell CD56dimCD16+ (p=1.5e-23)<br>CD4+ Th2 (p=4.4e-23)<br>Monocytes, classical. (p=9.6e-22)<br>CD4+ memory TREG (p=1.3e-21)<br>CD4+ Th17 (p=2.6e-21)<br>CD4+ Th1/Th17 (p=3.0e-21)<br>Monocyte, M2 (p=9.2e-21)<br>CD8+ naïve (p=8.7e-19)<br>CD4+ naïve (p=1.1e-18)<br>CD4+ Th1 (p=9.9e-18)<br>CD8+ naïve (activated) (p=1.5e-13)<br>B cell naïve (p=1.1e-12)<br>CD4+ naïve (activated) (p=9.0e-12) |
| “ |  | *CTD-2260A17.1*<br>(ENSG00000248734) | CD4+ naïve (p=0.00070) **T>A** |
| rs1820149 | Reference allele: T<br>Alternate allele: A | *ERAP2* | **A>T**<br>CD4+ naïve TREG (p=1.6e-24)<br>CD4+ TFH (p=6.6e-24)<br>NKcell CD56dimCD16+ (p=1.5e-23)<br>CD4+ Th2 (p=4.4e-23)<br>Monocytes, classical. (p=9.6e-22)<br>CD4+ memory TREG (p=1.3e-21)<br>CD4+ Th17 (p=2.6e-21)<br>CD4+ Th1/Th17 (p=3.0e-21)<br>Monocyte, M2 (p=9.2e-21)<br>CD8+ naïve (p=8.7e-19)<br>CD4+ naïve (p=1.1e-18)<br>CD4+ Th1 (p=9.9e-18)<br>CD8+ naïve (activated) (p=1.5e-13)<br>B cell naïve (p=1.1e-12) |
|  |  |  | CD4+ native (activated) (p=9.0e-12) |
| " |  | *CTD-2260A17.1*<br>(ENSG00000248734) | CD4+ naïve (p=0.00070) **A>T** |
| **SNP** |  | **Gene** | **Cells Impacted (adjusted p=values) – Allelic effects** |
| rs1559267 | Reference allele: A<br>Alternate allele: T | *ERAP2* | **T>A**<br>CD4+ naïve TREG (p=1.6e-24)<br>CD4+ TFH (p=6.6e-24)<br>NKcell CD56dimCD16+ (p=1.5e-23)<br>CD4+ Th2 (p=4.4e-23)<br>Monocytes, classical. (p=9.6e-22)<br>CD4+ memory TREG (p=1.3e-21)<br>CD4+ Th17 (p=2.6e-21)<br>CD4+ Th1/Th17 (p=3.0e-21)<br>Monocyte, M2 (p=9.2e-21)<br>CD8+ naïve (p=8.7e-19)<br>CD4+ naïve (p=1.1e-18)<br>CD4+ Th1 (p=9.9e-18)<br>CD8+ naïve (activated) (p=1.5e-13)<br>B cell naïve (p=1.1e-12)<br>CD4+ native (activated) (p=9.0e-12) |
| " |  | *CTD-2260A17.1*<br>(ENSG00000248734) | CD4+ naïve (p=0.00070) **T>A** |
| rs35675823 | Reference allele: C<br>Alternate allele: G | *CCR2* | **C>G**<br>CD4+ Th17 (p=2.6e-9)<br>CD4+ TFH (p=0.000046)<br>CD4+ Th1/Th17 (p=0.0032) |

We next sought to determine whether MPRA-identified SNPs were more likely to be DICE eQTLs than SNPs that screened negative (i.e., did not show allele-specific differences) on the MPRA. To do this, we used the “randomize” function in the Excel spreadsheet that was used to create the oligo library (the SNPs were ordered by chromosome and chromosome position) and selected SNPs sequentially from the re-ordered column. We excluded SNPs that were in the major histocompatibility complex (MHC-Chr6:30,206,688-32,920,187), as these SNPs were also excluded in the original analysis (they were included on the original MPRA as part of a project examining the role of non-coding regulatory functions within the MHC, which is ongoing). We found that randomly selected SNPs were less likely to be eQTLs in DICE than the n=84 that did (38/84 from the MPRA vs 60/200 from the randomly selected set; Chi-squared with Yates’ continuity correction = 5.42; p=0.0199; OR=1.93, 95% CI=2.8-27.6).

Although the MPRA was performed in K562 cells, a lymphoblastic cell line derived from a patient with chronic myelogenous leukemia, gene expression effects associated with MPRA-detected SNPs were associated with gene expression levels across a range of human immune cell types and subtypes. This was predictable, as hematopoietic cells demonstrate broad commonalities in transcription factor expression and structures of transcriptional networks(22). This was particularly true for SNPs within the *ERAP2/LNPEP* locus. For example, rs1216565, is associated with significant allele-specific effects on transcription in a range of CD4+ T cell subsets, as well as CD8+ T cells, NK cells, and B cells. These findings are illustrated in **Figure 1**. Similarly, rs1194608, which is associated with allele-specific transcription levels of the gene, *ATP8B2* within the *IL6R/ATP8B2* locus, exerts its effects in both CD8+ and CD4+ T cells, as illustrated in **Figure 2**.

**Figure 1.**
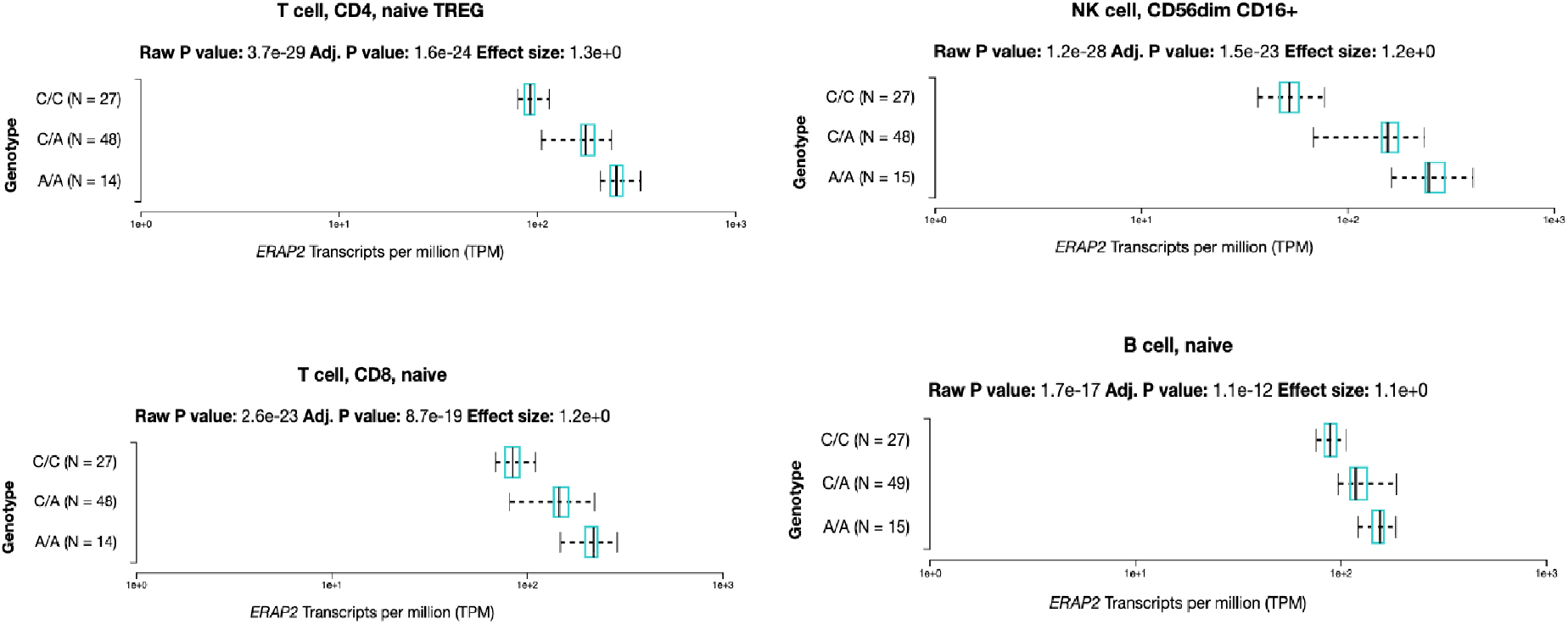
Log-scale bar charts derived from Database of Immune Cell eQTL analyses of transcriptional effects of rs1216565, a SNP within the juvenile arthritis-associated *ERAP2* locus. Significant allele-specific effects of this SNP are identified across a range of immune cells, with individuals homozygous for the A allele showing significantly higher expression of *ERAP2* than individuals who are homozygous for the C allele. Heterozygotes show intermediate levels of expression.

**Figure 2.**
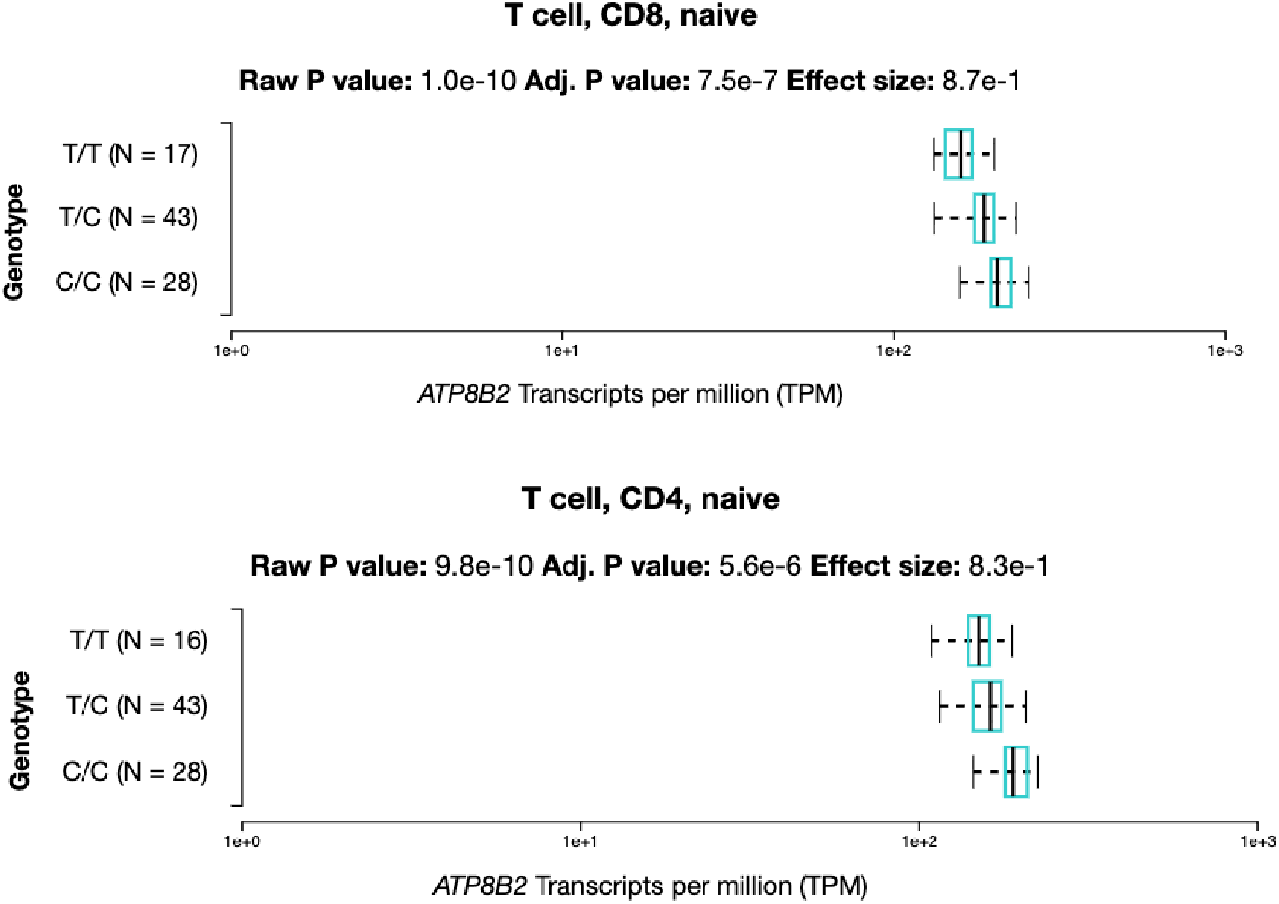
Log-scale bar charts derived from Database of Immune Cell eQTL analyses of transcriptional effects of rs1194608, a SNP within the juvenile arthritis-associated risk locus, *IL6R/ATP8B2*. Significant allele-specific effects of this SNP are identified in both naïve CD8+ and naïve CD4+ T cells, with individuals homozygous for the C allele showing significantly higher expression of *ATP8B2* than individuals who are homozygous for the T allele. Heterozygotes show intermediate levels of expression.

The finding that multiple SNPs are associated with alterations in gene expression in CD8+ T cells adds further evidence to the importance of CD8+ T cells in JIA, a feature of the immunopathology of the disease that is being increasingly recognized(23–25). Another interesting feature that emerges from the analysis of the DICE data is the apparent genetic influences of the expression of long non-coding RNAs, which we have previously identified within the JIA risk regions from neutrophil RNAseq data(4). The findings from the *TRAF1* locus are therefore of some interest. For example, rs1014529 (**Table 4**) is a SNP within an H3K27ac-marked intronic region within the *TRAF1* gene and is associated with allelic effects on expression of *TRAF1* in CD4+ T follicular helper cells, and both naïve and memory regulatory T cells (TREG). At the same time, this SNP is associated with transcription effects across a broad range of CD4+ T cell subtypes, CD8+ T cells subtypes, B cells, and monocytes for *PSMD5-AS1* (ENSG00000226752), as illustrated in **Figure 3**. *PSMD5-AS1* is a long non-coding RNA encoded on the anti-sense strand of the *PSMD5* gene, >100,000 bp from the *TRAF1* transcription start site but within the same topologically associated domain in numerous immune cell types(21). *PSMD5-AS1* has recently been identified as a risk gene for JIA in both European and Asian ancestries by genome-wide transcriptome analysis(26). Other non-coding RNAs whose expression levels are associated with MPRA-identified SNPs are shown in **Tables 4** and **5**. For clarity, these transcripts are also identified by their ENSEMBL gene names.

**Figure 3.**
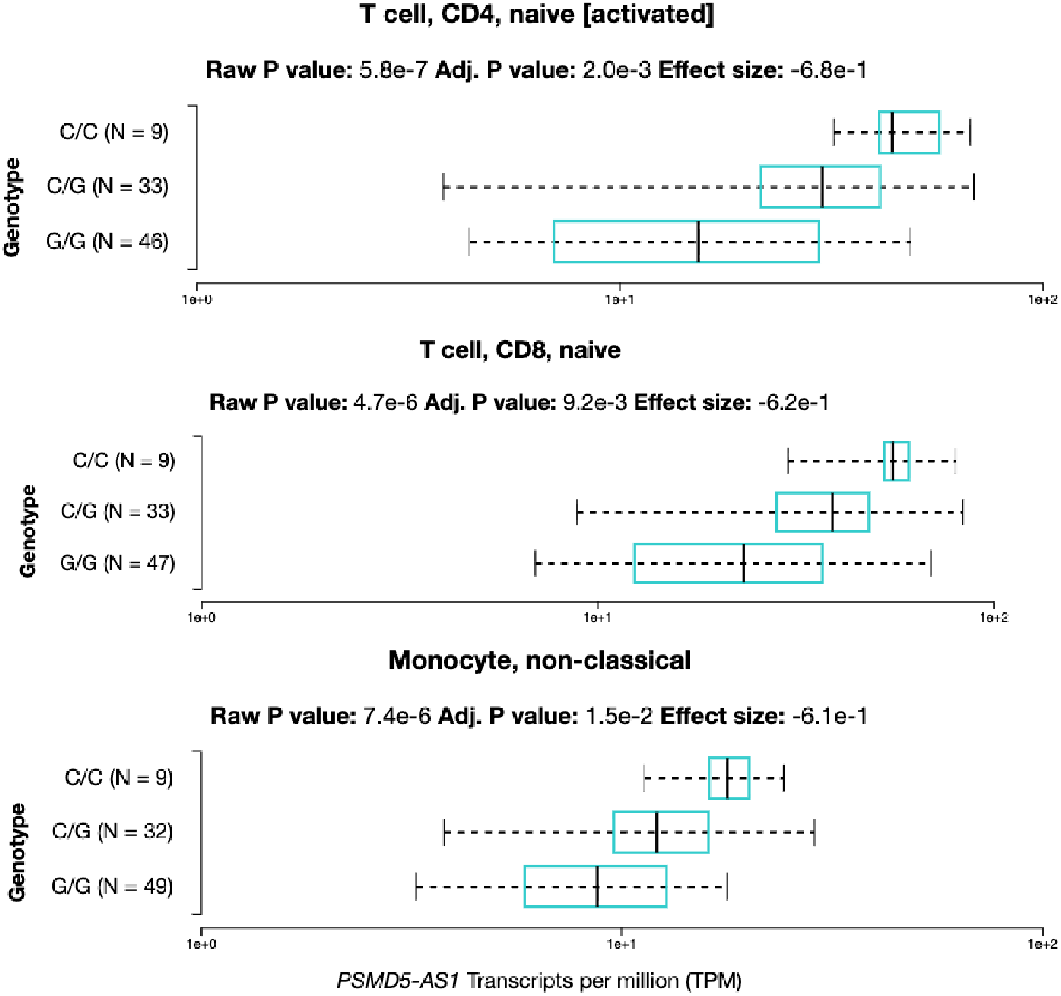
rs1014529, a SNP situated within a *TRAF1* H3K27ac-marked intronic region, is associated with transcriptional effects on *PSMD5-AS1*, a non-coding RNA encoded on the antisense strand of the gene, *PSMD5,* situated ∼100,000 bp from the *TRAF1* transcription start site, but within the same topologically associated domain (TAD) (21). Log-scale bar charts are derived from the Database for Immune Cell eQTLs and show transcription effects on a broad range of immune cell types (see also **Table 3**).

**Table 5:**
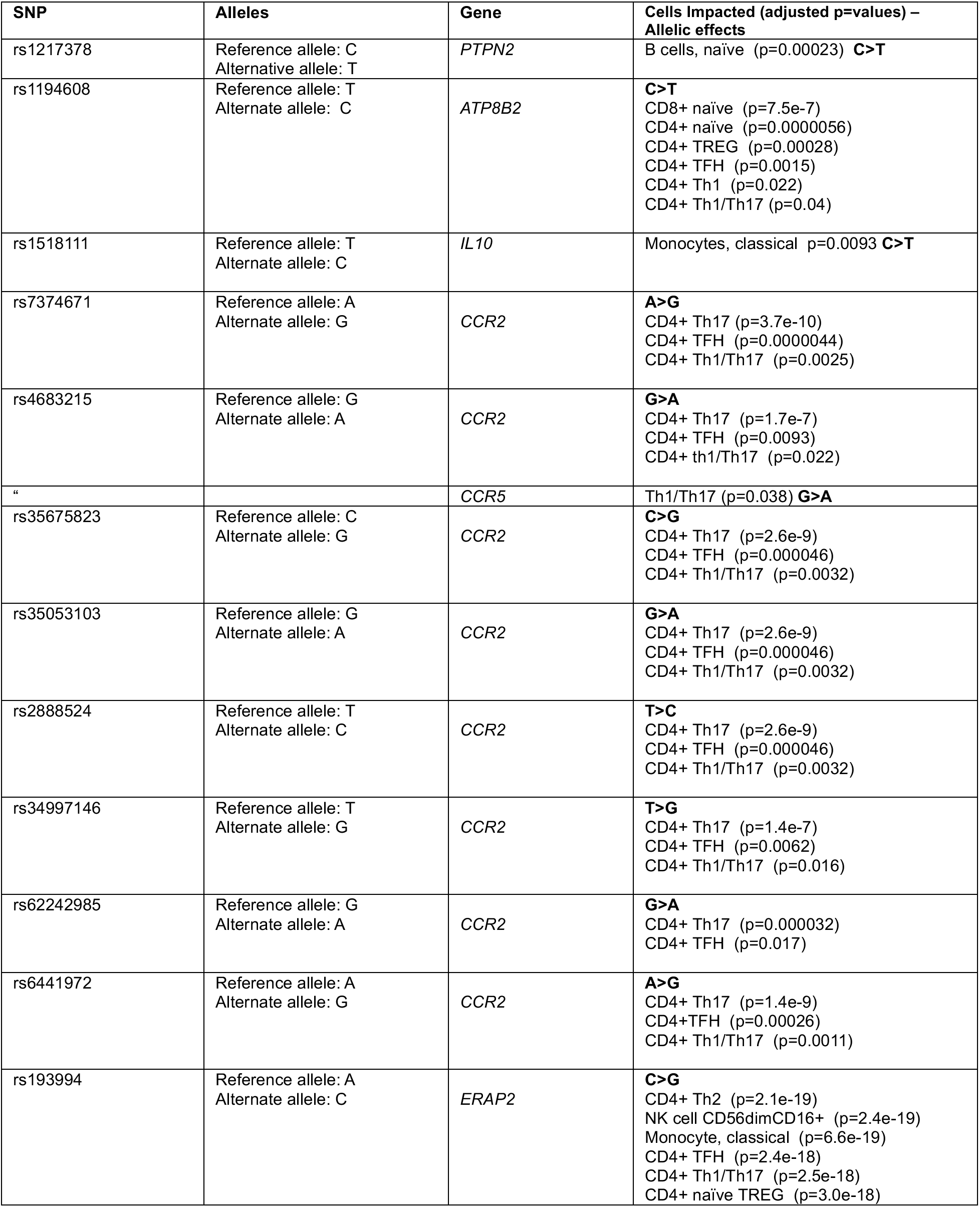
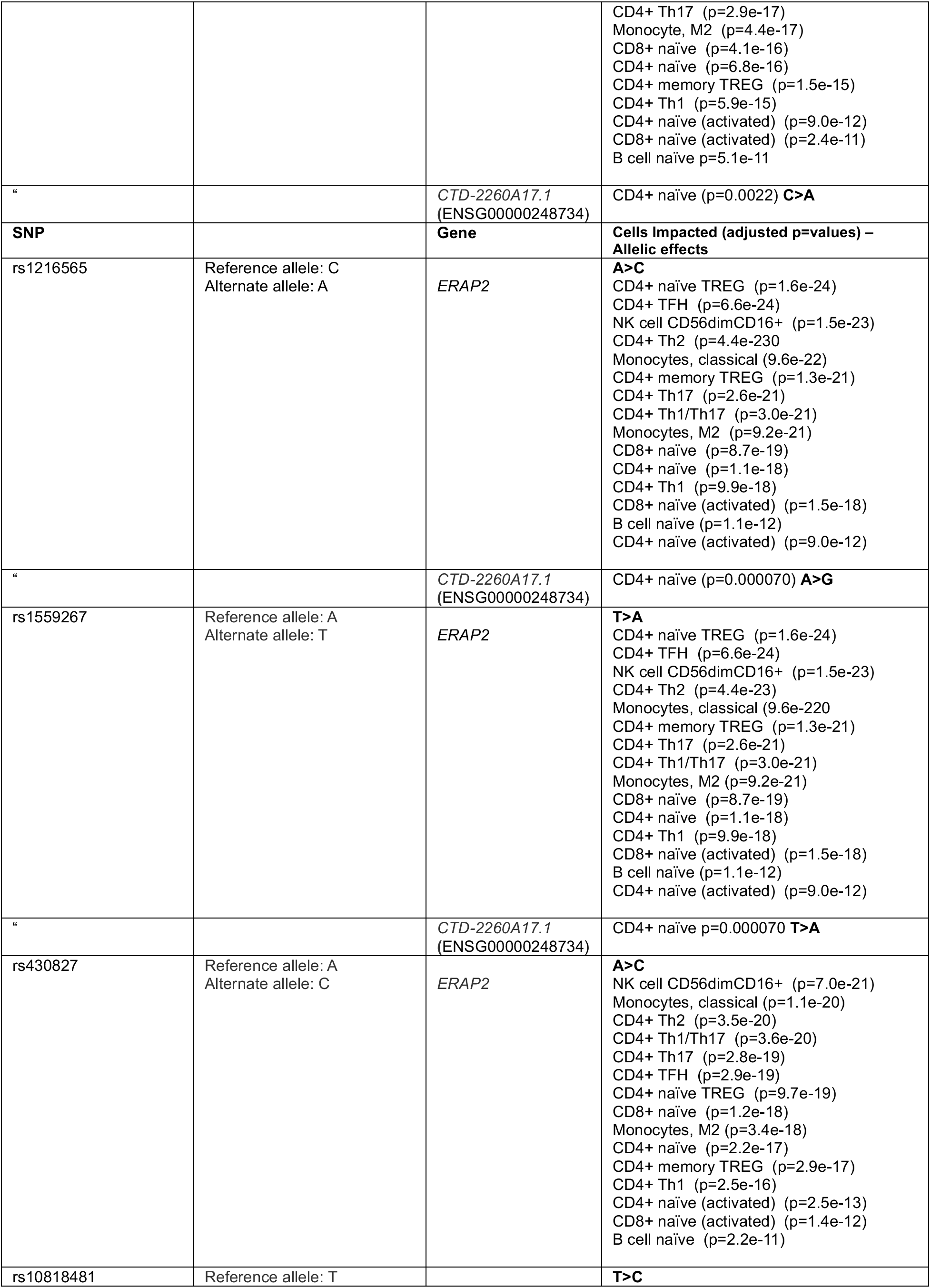
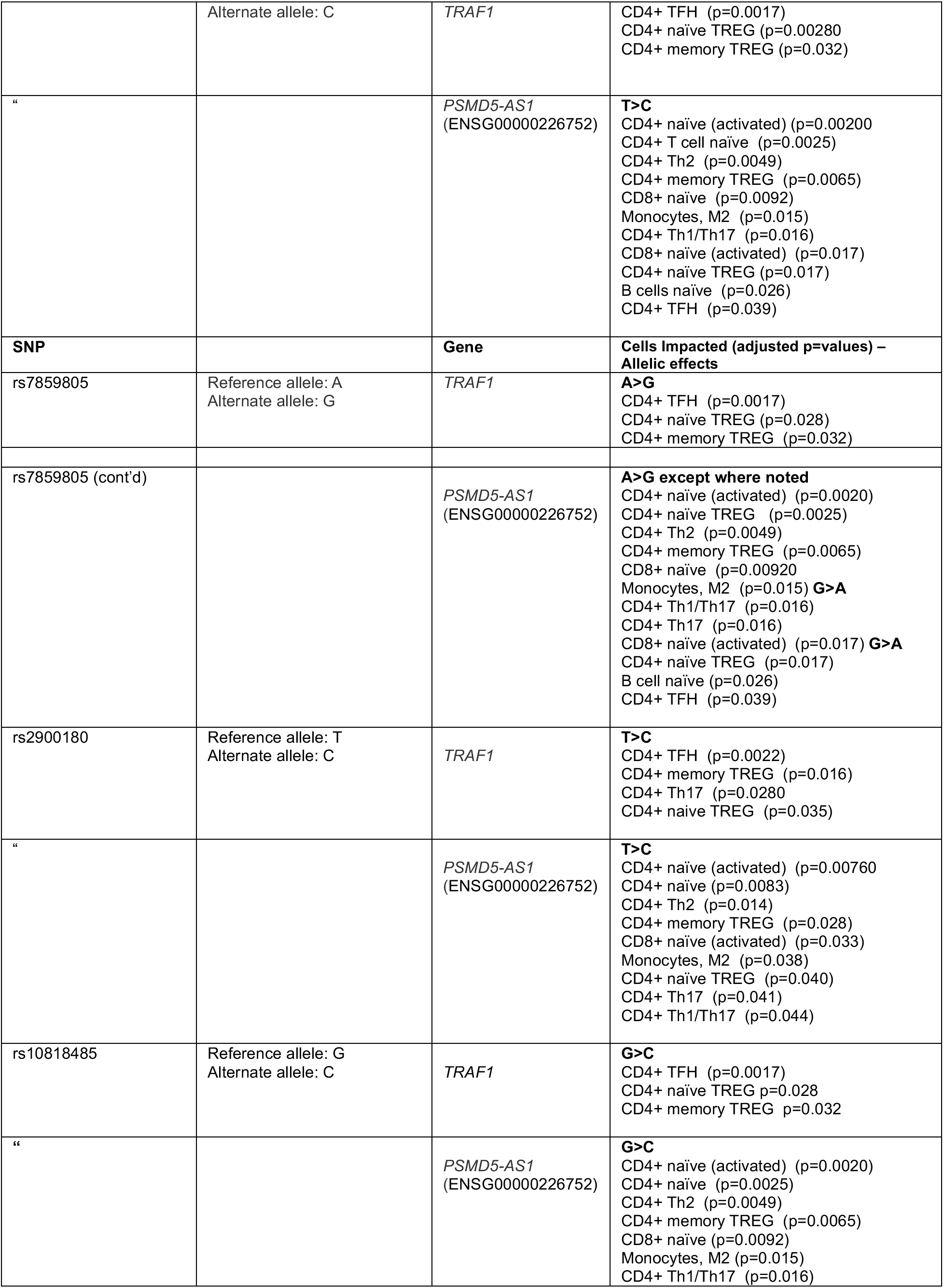
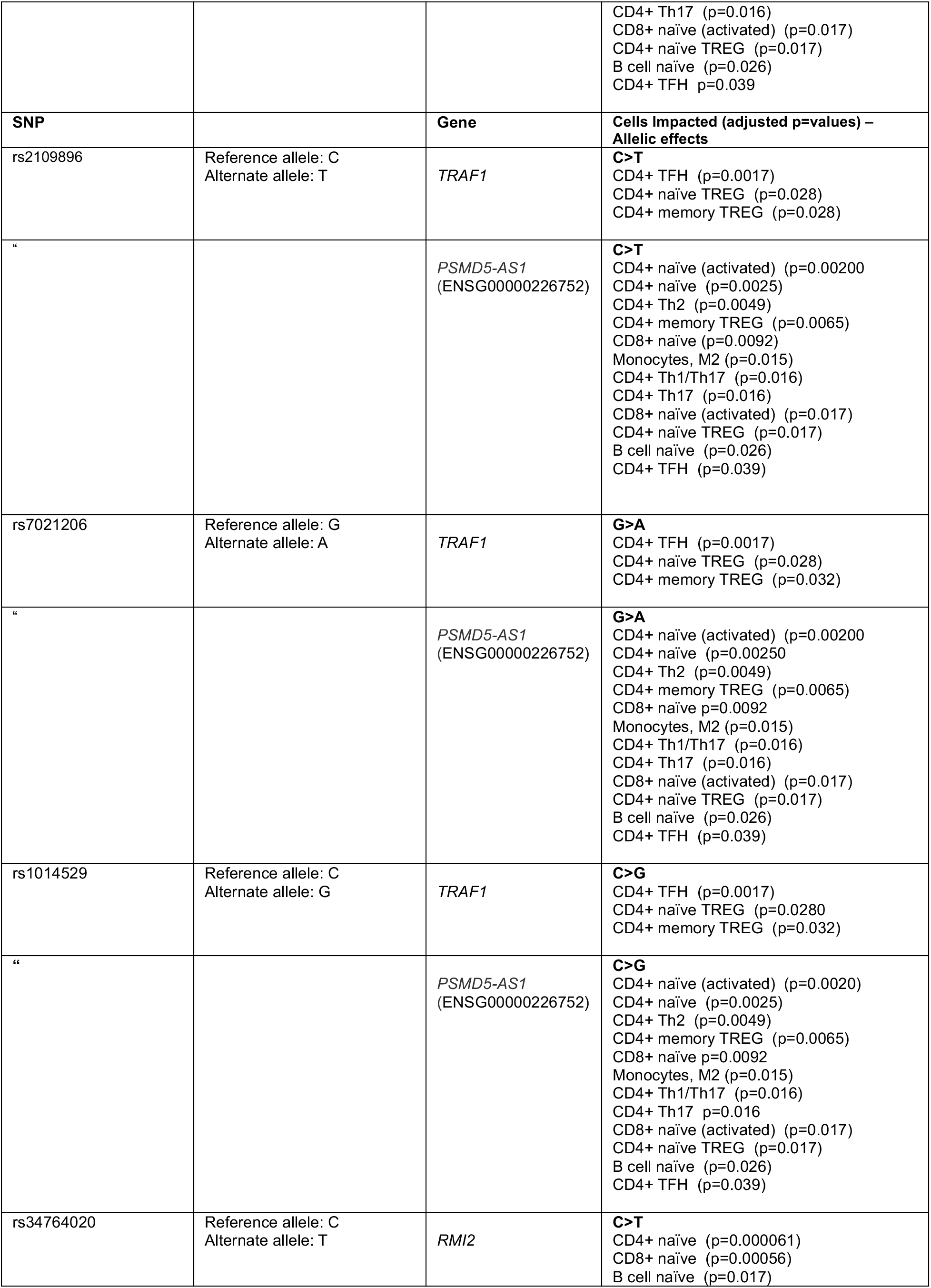
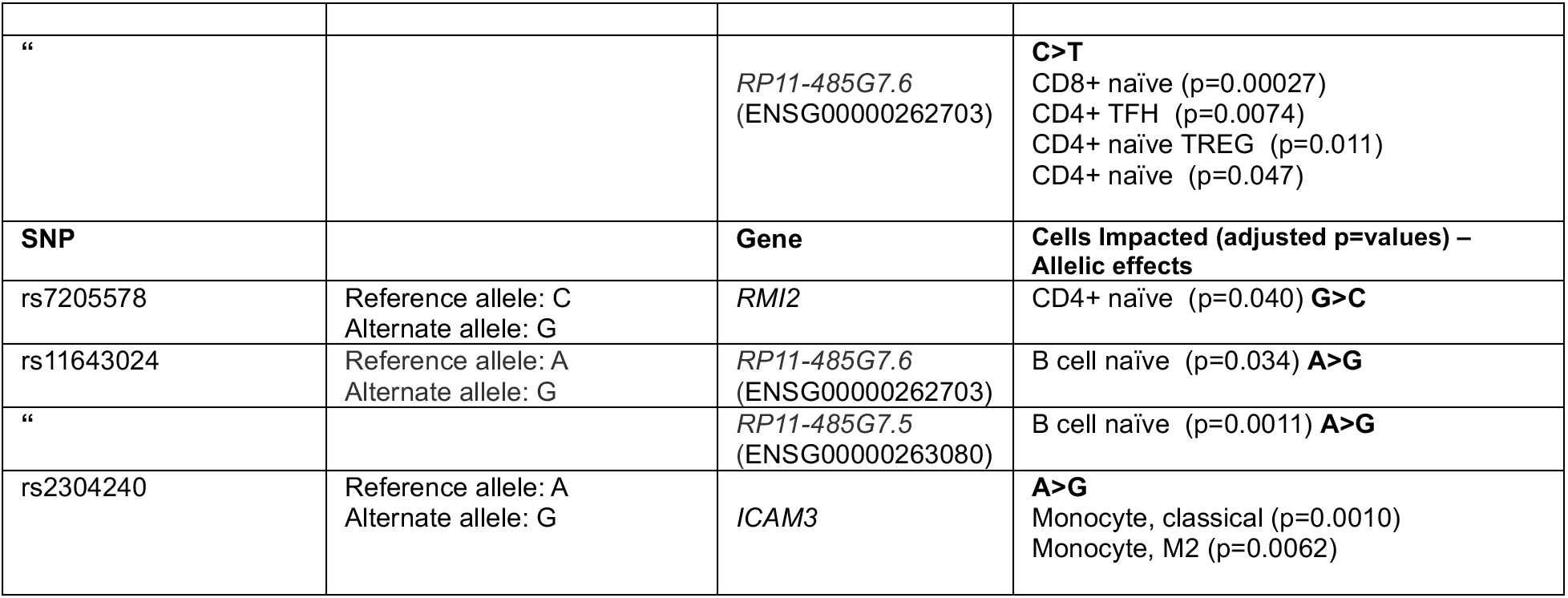
DICE eQTLs for MPRA-Screened SNPs: IFN gamma Stimulated K562 Cells. Abbreviations: TFH – T follicular helper cells TREG – Regulatory T cells NK – Natural Killer

### 3.2. Analyses From GTEx

A significant shortcoming of the DICE data is that they were generated from a small number of individuals. Thus, the DICE data are likely underpowered to detect small but meaningful genetic impacts on transcription or to assess transcriptional effects of less-abundant alleles. We therefore used the Gene-Tissue Expression (GTEx) project data from whole blood, EBV-transformed lymphocytes, and spleen to determine the extent to which MPRA-identified SNPs are associated with expression of relevant genes within the GTEx project tissues. These data are summarized in **Tables 6** and **7**. Among the n=42 SNPs identified in unstimulated K562 cells, n=25 (60%) were associated with gene expression levels in one or more GTEx tissues. Among the n=42 SNPs identified in K562 cells stimulated with IFNγ, n=40 (95%). were associated with gene expression levels in one or more of the interrogated GTEx tissues. Many of the SNPs (n=38) that were associated with levels of gene expression in the GTEx tissues were not detected in the DICE data. For those SNPs that were eQTLs in both DICE and GTEx data, the was strong concordance in the allele identified as being associated with the higher levels of expression of the identified cells/target tissues. The exception was rs11643024, within the *RMI2* risk region. In DICE data, this SNP was identified as being associated with expression of the lncRNA, *RP11-485G7*.6 *(ENSG00000262703*) in B cells, with individuals homozygous for the reference A allele showing higher expression that individuals homozygous for the alternate G allele. In the GTEx data, this SNP is associated with expression levels of *RMI2* in EBV-transformed lymphocytes, with individuals homozygous for the alternate allele (G) showing higher expression than individuals homozygous for the reference A allele.

**Table 6:**
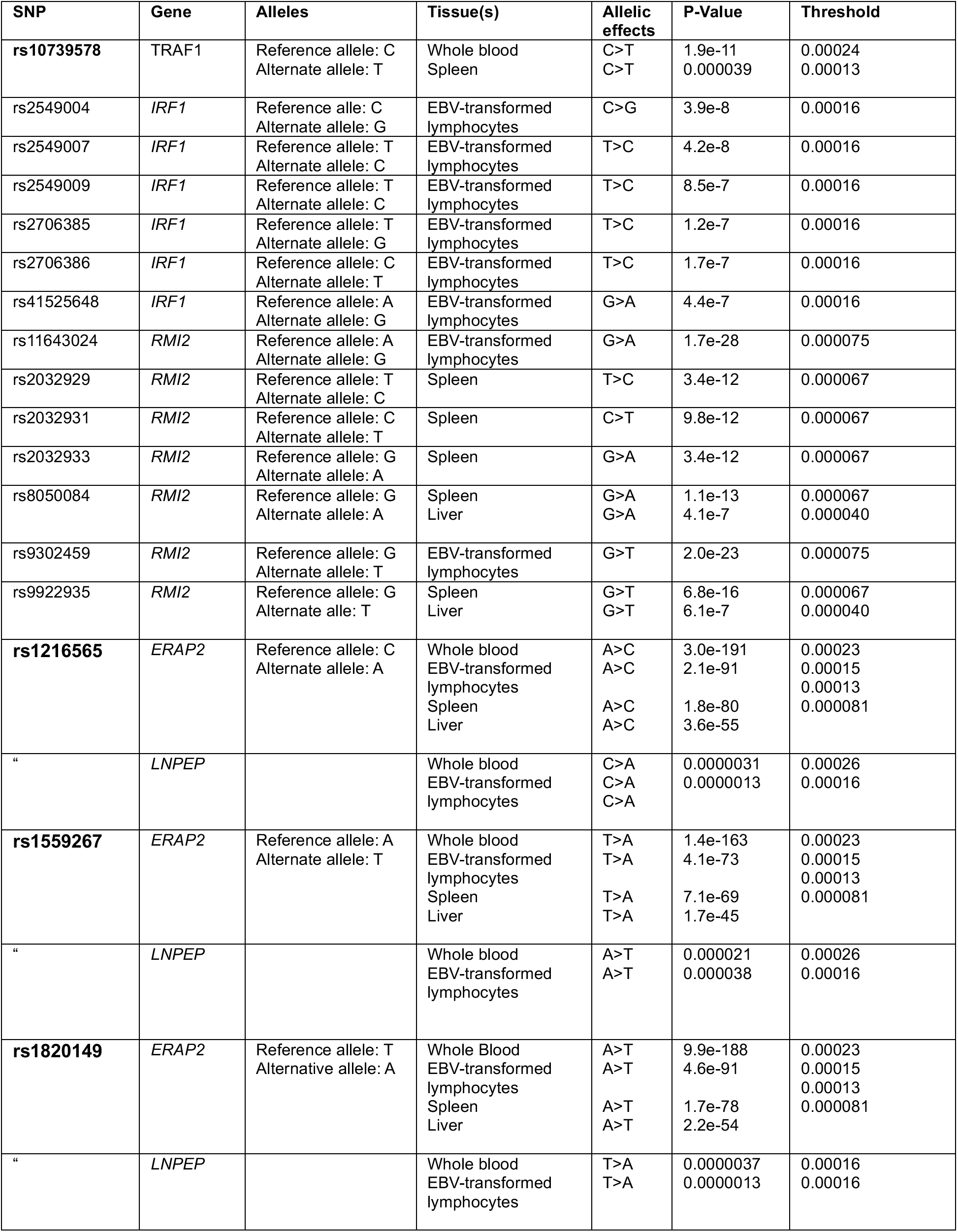
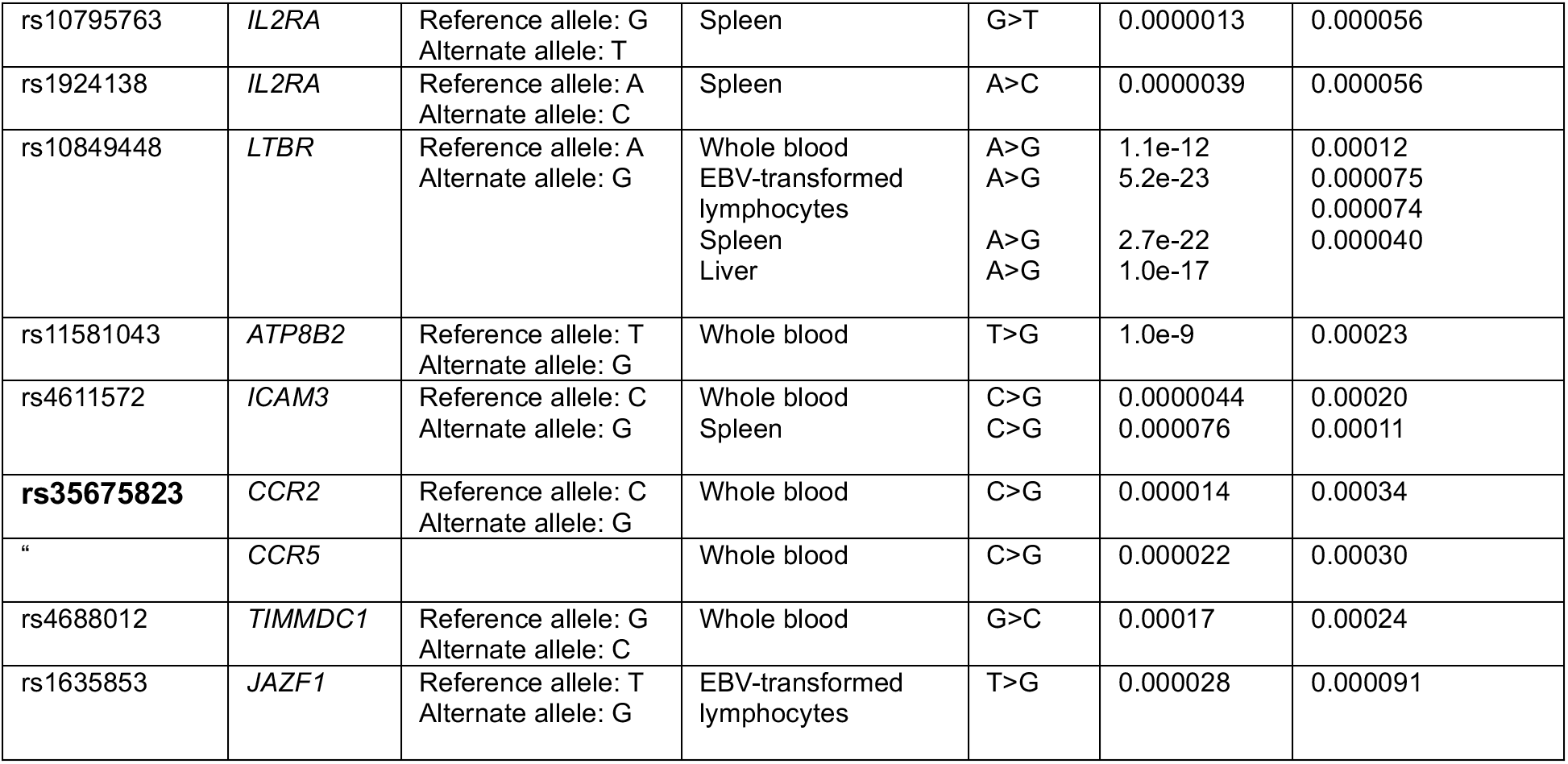
Unstimulated K562 SNPs With GTEx Data (SNPs in bold type also have DICE effects)

**Table 7:** K562 + IFNg SNPs With GTEx Data (SNPs in bold also have DICE effects)

| SNP | Gene | Alleles | Tissue(s) | Allelic Effects | p-Value | Threshold |
| --- | --- | --- | --- | --- | --- | --- |
| rs11581043 | *ATP8B2* | Reference allele:<br>T<br>Alternate allele:<br>G | Whole blood | G>T | 1.0e-9 | 0.00023 |
| **rs1518111** | *IL10* | Reference allele:<br>T<br>Alternate allele:<br>C | Whole blood | C>T | 1.0e-22 | 0.00019 |
| **rs1217378** | *PTPN22* | Reference allele:<br>C<br>Alternate allele:<br>T | Spleen | C>T | 0.0000029 | 0.00013 |
| rs2177812 | *TIMMDC1* | Reference allele:<br>G<br>Alternate allele:<br>T | Whole blood | G>T | 0.00017 | 0.00034 |
| **rs193994** | *ERAP2* | Reference allele:<br>A<br>Alternate allele:<br>C | Whole blood<br>EBV-transformed lymphocytes<br>Spleen<br>Liver | C>A<br>C>A<br>C>A<br>C>A | 2.5e-185<br>2.7e-85<br>1.7e-77<br>1.7e-57 | 0.00023<br>0.00015<br>0.00013<br>0.000081 |
| “ | *LNPEP* |  | Whole blood<br>EBV-transformed lymphocytes | A>C<br>A>C | 0.0000030<br>3.5e-7 | 0.00026<br>0.00016 |
| **rs1216565** | *ERAP2* | Reference allele:<br>C<br>Alternate allele:<br>A | Whole blood<br>EBV-transformed lymphocytes<br>Spleen<br>Liver | A>C<br>A>C<br>A>C<br>A>C | 3.0e-191<br>2.8e-91<br>1.8e-80<br>3.5e-55 | 0.00023<br>0.00015<br>0.00013<br>0.000081 |
| “ | *LNPEP* |  | Whole blood<br>EBV-transformed lymphocytes | C>A<br>C>A | 0.0000031<br>0.0000013 | 0.00026<br>0.00016 |
| **rs1559267** | *ERAP2* | Reference allele:<br>A<br>Alternate allele:<br>T | Whole blood<br>EBV-transformed lymphocytes<br>Spleen<br>Liver | T>A<br>T>A<br>T>A<br>T>A | 1.4e-163<br>4.1e-73<br>7.1e-69<br>1.7e-45 | 0.00023<br>0.00015<br>0.00013<br>0.000081 |
| “ | *LNPEP* |  | Whole blood<br>EBV-transformed lymphocytes | A>T<br>A>T | 0.000021<br>0.000038 | 0.00026<br>0.00026 |
| **rs430827** | *ERAP2* | Reference allele:<br>C<br>Alternate allele:<br>A | Whole blood<br>EBV-transformed lymphocytes<br>Spleen<br>Liver | A>C<br>A>C<br>A>C<br>A>C | 1.8e-267<br>2.3e-118<br>1.4e-90<br>5.6e-82 | 0.00023<br>0.00015<br>0.00013<br>0.000081 |
| “ | *LNPEP* |  | Whole blood<br>EBV-transformed lymphocytes | A>C<br>C>A | 6.2e-11<br>0.0000071 | 0.00026<br>0.00016 |
| rs41525648 | *IRF1* | Reference allele:<br>A<br>Alternate allele:<br>G | EBV-transformed lymphocytes | G>A | 4.4e-7 | 0.00016 |

| SNP | Gene |  | Tissue |  | p-Value | Threshold |
| --- | --- | --- | --- | --- | --- | --- |
| rs2706385 | *IRF1* | Reference allele:<br>T<br>Alternate allele:<br>G | EBV-transformed lymphocytes | G>T | 1.2e-7 | 0.00016 |
| rs2706386 | *IRF1* | Reference allele:<br>C<br>Alternate allele:<br>T | EBV-transformed lymphocytes | T>C | 1.7e-7 | 0.00016 |
| rs1924138 | *IL2RA* | Reference allele:<br>A<br>Alternate allele:<br>C | Spleen | A>C | 0.0000039 | 0.000056 |
| rs10795763 | *IL2RA* | Reference allele:<br>G<br>Alternate allele:<br>T | Spleen | G>T | 0.0000013 | 0.00056 |
| rs8050084 | *RMI2* | Reference allele:<br>G<br>Alternate allele:<br>A | Spleen | G>A | 1.1e-14 | 0.000067 |
| rs9302459 | *RMI2* | Reference allele:<br>G<br>Alternate allele:<br>T | EBV-transformed lymphocytes | G>T | 2.0e-23 | 0.000075 |
| rs6498184 | *RMI2* | Reference allele:<br>T<br>Alternate allele:<br>C | Spleen | T>C | 5.6e-17 | 0.000067 |
| **rs11643024** | *RMI2\** | Reference allele:<br>A<br>Alternate allele:<br>G | EBV-transformed lymphocytes | G>A | 1.7e-28 | 0.000067 |
| rs2032929 | *RMI2* | Reference allele:<br>T<br>Alternate allele:<br>C | Spleen | T>C | 3.4e-12 | 0.000067 |
| rs2032931 | *RMI2* | Reference allele:<br>C<br>Alternate allele:<br>T | Spleen | C>T | 9.8e-12 | 0.000067 |
| rs2032933 | *RMI2* | Reference allele:<br>G<br>Alternative allele:<br>A | Spleen | G>A | 3.4e-12 | 0.000067 |
| rs9952753 | *PTPN2* | Reference allele:<br>A<br>Alternate allele:<br>C | EBV-transformed lymphocytes | C>A | 0.0000059 | 0.000089 |
| rs515151 | *PTPN2* | Reference allele:<br>G<br>Alternate allele:<br>C | Spleen | G>C | 0.0000031 | 0.000099 |
| **rs2304240** | *ICAM3* | Reference allele:<br>A<br>Alternate allele:<br>G | Whole blood.<br>Spleen | A>G<br>A>G | 8.2e-16<br>0.0000052 | 0.00020<br>0.00011 |
| rs4611572 | *ICAM3* | Reference allele:<br>C<br>Alternate allele:<br>G | Whole blood<br>Spleen | C>G<br>C>G | 0.0000044<br>0.000076 | 0.00020<br>0.00011 |
| rs510506 | *ICAM3* | Reference allele:<br>G<br>Alternate allele:<br>T | Whole blood | G>T | 1.3e-11 | 0.00020 |
| **rs7374671** | *CCR2* | Reference allele:<br>A | Whole blood | A>G | 0.000036 | 0.00034 |

|  |  | Alternate allele:<br>G |  |  |  |  |
| --- | --- | --- | --- | --- | --- | --- |
| SNP | Gene | Alleles | Tissue(s) | Allelic Effects | p-Value | Threshold |
| **rs4683215** | *CCR2* | Reference allele:<br>G<br>Alternate allele:<br>A | Whole blood | G>A | 2.0e-9 | 0.00034 |
| “ | *CCR5* |  | Whole blood | G>A | 0.000041 | 0.00030 |
| **rs35675823** | *CCR2* | Reference allele:<br>C<br>Alternate allele:<br>G | Whole blood | C>G | 0.000014 | 0.00034 |
| “ | *CCR5* |  | Whole blood | C>G | 0.000024 | 0.00030 |
| **rs35053103** | *CCR2* | Reference allele:<br>G<br>Alternate allele:<br>A | Whole blood | G>A | 0.0000077 | 0.00034 |
| “ | *CCR5* |  | Whole blood | G>A | 0.000027 | 0.00030 |
| rs2888523 | *CCR2* | Reference allele: T<br>Alternate allele:<br>C | Whole blood | T>C | 9.7e-9 | 0.00034 |
| **rs2888524** | *CCR2* | Reference allele:<br>T<br>Alternate allele:<br>C | Whole blood | T>C | 0.000011 | 0.00034 |
| “ | *CCR5* |  | Whole blood | T>C | 0.000017 | 0.00030 |
| **rs34997146** | *CCR2* | Reference allele:<br>T<br>Alternate allele:<br>G | Whole blood | T>G | 2.0e-9 | 0.00034 |
| “ | *CCR5* |  | Whole blood | T>G | 0.000041 | 0.00030 |
| **rs62242985** | *CCR2* | Reference allele:<br>G<br>Alternate allele:<br>A | Whole blood | G>A | 0.000055 | 0.00034 |
| **rs6441972** | *CCR5* | Reference allele:<br>A<br>Alternate allele:<br>G | Whole blood | A>G | 0.00012 | 0.00030 |

\*In DICE data, the SNP is associated with expressions levels of the ncRNA, *RP11-485G7.6* (ENSG00000262703), in B cells, with A>G.

It is also worth noting that, given that the GTEx whole blood expression data reflect a strong neutrophil signature(27), a feature we have also detected in whole blood expression data from children with JIA(28, 29), these analyses provide indirect evidence that risk-driving genetic variants for JIA may influence neutrophils, as we have previously suggested(4, 30). Genetic influences on gene expression were detected in whole blood for 14 of 42 SNPs detected by MPRA in unstimulated K562 cells (33%) and in 22 of the 42 SNPs (52%) detected in IFNγ-stimulated K562 cells.

Together, the DICE and GTEx data also provide useful insights into the like target genes, and therefore affected immune pathways, in JIA. For example, genetic variants in the *TYK2* region, including variants within the coding sequences, are associated with a broad range of autoimmune diseases(31), including JIA. Since TYK2 is an important tyrosine kinase that mediates the downstream effects of type 1 interferons, there is considerable interest in TYK2 as a therapeutic target. However, neither the DICE nor the GTEx data identified *TYK2* as an affected gene in JIA. Our data do support the idea that *ICAM3*, an important cell-surface molecule for T cell adhesion and activation(32), is a gene upon which genetic risk is exerted. It’s worth noting that ICAM3 also plays an important role in myeloid cells(33), and the DICE and GTEx data support the idea that genetic effects are exerted on lymphoid and myeloid cells. **Figure 4** shows DICE eQTL data for rs2304240, which, in DICE data, exerts effects on *ICAM3* levels exclusively in myeloid cells. None of these findings exclude the possibility that genetic influences that drive JIA risk are exerted on *TYK2* expression. However, they do provide evidence that genetic risk may also be exerted on other genes on the TYK2 haplotype.

**Figure 4.**
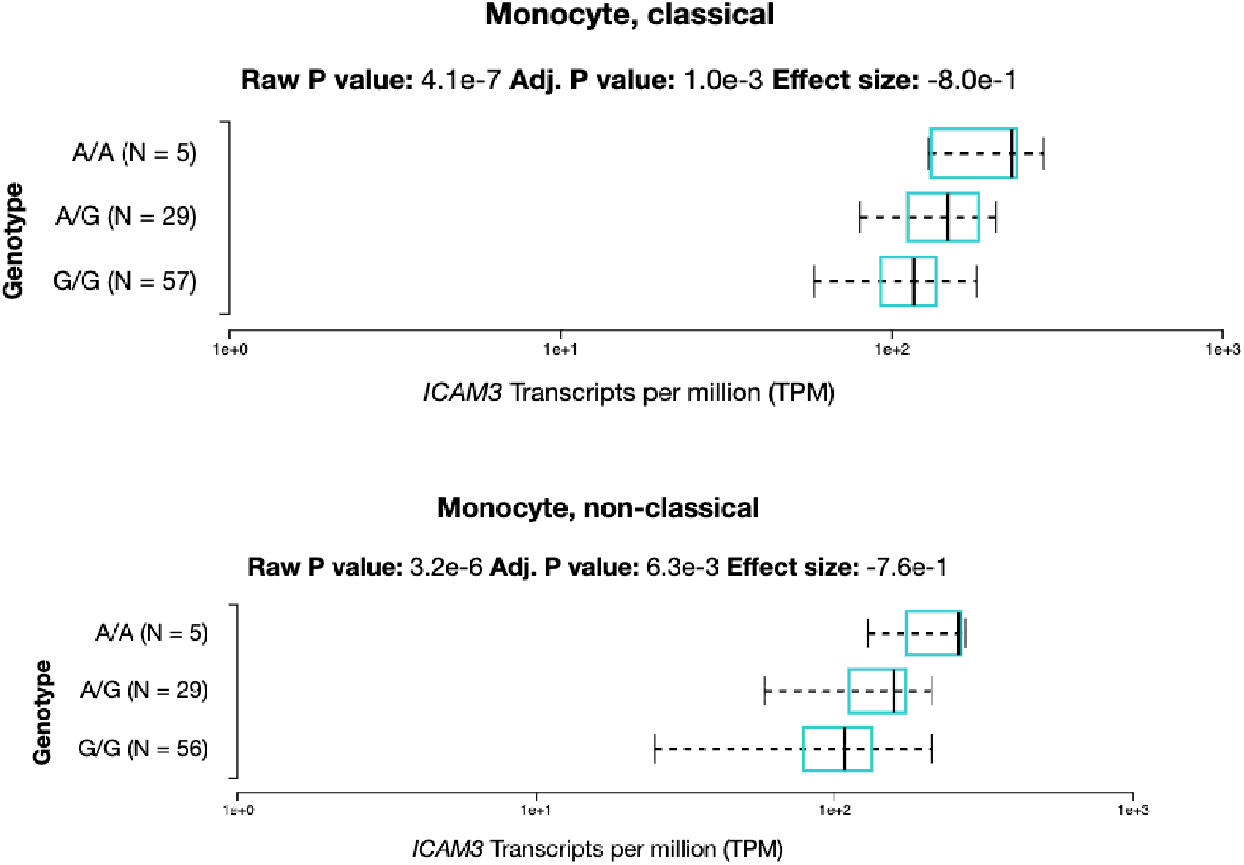
Log-scale bar charts derived from Database of Immune Cell eQTL analyses of transcriptional effects of rs2304240, a SNP within the juvenile arthritis-associated risk locus, *TYK2/ICAM3*. Significant allele-specific effects of this SNP are identified in both classical and non-classical (M2) monocytes, with individuals homozygous for the A allele showing significantly higher expression of *ICAM3* than individuals who are homozygous for the G allele. Heterozygotes show intermediate levels of expression.

## 4. Discussion

MPRA are a widely-used tool for screening SNPs on autoimmune disease risk haplotypes for a pathologically-relevant feature: the impact of specific alleles on gene expression (12, 34–36). Selection of this feature as a focus of screening is supported by the abundance of evidence demonstrating that genetic risk for autoimmunity is most likely to impact cis-regulatory elements of disease-influencing genes rather than alterations in the coding sequences(4, 10). There are, however, aspects of the design of MPRA that may limit their applicability to human disease genetics. The first is the fact that MPRA don’t query SNPs/alleles in their actual genomic context. The 200 bp oligos therefore may lack complex 3D structures/shape(1) that are biologically and pathologically important. Next, MPRA are both relative expensive and labor-intensive to perform, with each step requiring focused quality-control measures before proceeding to the next(14). As a consequence, MPRA are typically performed in a single cell type, whereas we know that, for diseases like JIA, there is strong evidence that genetic risk is exerted across a range of immune cell types (30). In this paper we demonstrate that MPRA, even when performed in a human cell line, can used in combination with human gene expression data to elucidate a range of cells/tissues that may be impacted by genetic variants on JIA risk haplotypes as well as the genes whose expression levels are likely to be impacted by risk-driving variants on the JIA risk haplotypes. An important finding from this study is the broad range of cells likely to be impacted by JIA risk variants. We present evidence that CD4+ and CD8+ T cells and T cell subsets, B cells, monocytes, and NK cells may all be impacted by JIA risk-driving variants, consistent with our earlier findings (30). These conclusions can be drawn from MPRA even in a single cell type because of the cellular mechanism through which MPRA work. In MPRA, expression of the GFP reporter is driven by the panoply of transcription factors (TFs) within the host cell. Changes in GFP expression are driven by changes in the efficiency of TF binding to the oligos’ minimal promoter. Because leukocytes express a wide range of common TFs, it is therefore not surprising that an allele that disrupts a TF binding site in one cell type would do so in another, and that these effects would be reflected in expression levels of affected genes in those cells. Taken together, our findings support the idea of complex interactions between cells of the adaptive and innate immune systems including neutrophils(37) driving both genetic risk and pathobiology in JIA.

MPRA are agnostic with respect to an important issue: the identification of target genes. Because of the 3D character of the genome within the nucleus, it cannot be assumed that the gene affected by a risk-driving variant is the one whose transcription start site is most proximal to the SNP of interest. For example, we present evidence (**Figure 3**) that rs1014529, which is situated within an intronic region of the *TRAF1* gene, influences expression levels of *PSMD5-AS1* (ENSG00000226752), a long non-coding (lnc)RNA that lies >100,000 bp away from the *TRAF1* start site, but is within the same topologically associated domain. Similarly, it may be tempting to assume that *IL6R* is the affected gene within the *IL6R/ATP8B2* locus, given the known importance of *IL6R* in joint pathobiology of JIA (38). However, both the GTEx data (**Tables 5** and **6**) and the DICE data (**Table 4**) implicate *ATP8B2*, a gene encoding a phospholipid flippase, as the target of expression-altering alleles on the JIA risk haplotypes. *ATP8B2* is expressed in a broad spectrum of lymphoid cells, and highest in CD4+ T cells and CD4+ T cell subsets, although its function and impact on immune pathobiology remain unclear at this time. These findings do not exclude *IL6R* as a target gene influenced by risk-driving SNPs on that haplotype. Rather, they provide evidence that other genes on the same risk haplotype may also be influenced.

There are several limitations to this study that need to be acknowledged. The foremost is the fact that neither the DICE nor GTEx data disentangle LD. The relationship between a given SNP and expression levels of any gene in these databases is simply an association, an indication that individuals who carry one or another allele of a particular SNP have significantly different levels of expression of a specific gene in specific cells/tissues. It does not indicate that the queried SNP drives those differences. However, the MPRA is designed to assist in disentangling LD, as it measures a property inherent in the tested SNPs: the intrinsic capacity of one allele or the other to drive differences in gene expression. Thus, MPRA strengthens the idea that the associations derived from GTEx and DICE data may be more than associations.

A curious finding is that K562 MPRA shows identifies multiple SNPs on the same haplotype that show intrinsic, allele-specific effects on expression of the GFP reporter (we have seen this in other MPRAs we have performed in primary human cells as well). Notable regions in the include the *ERAP2/LNPEP, RMI2,* and *CCR2* loci. The idea that there may be more than one SNP on the same haplotype driving risk for complex traits is becoming increasingly accepted(39, 40). However, one has to interpret the MPRA data cautiously and recognize their limitations. The strength of MPRA is their capacity to screen tens of thousands of variants for a biological property that is disease-relevant: the intrinsic capacity of a variant (within a somewhat artificial system) to influence gene expression. The MPRA do not identify the “risk-driving variants.” Rather, they screen and nominate candidate variants for further prioritization and characterization. Thus, it is unlikely that each of the MPRA-identified SNPs within the *ERAP2/LNPEP, RMI2,* and *CCR2* loci exerts independent effects.

DICE and GTEx data also have individual weaknesses for dissection genetic mechanisms driving risk. DICE data are limited in their ability to identify eQTLs by the relatively small number (n=91) of individuals in the data set. Thus, false negatives (i.e., failure to identify an association between a SNP and gene/transcript expression levels) are likely if relying solely on DICE data. Similarly, one must cautiously interpret the apparent cell-type specificity of the DICE data. Differences in sample size, differences in the abundance of specific cell types in individuals, allele frequency, and expression levels all limit our ability to make decisive conclusions with respect to the range of impacted cells The GTEx data set, in contrast, sampled 948 post-mortem donors, which may explain why more of the MPRA-identified SNPs could be identified as eQTLs in GTEx than in the DICE data. Furthermore, the GTEx data set includes whole blood, which, as previously noted, reflects a strong neutrophil signature(27) and thus may help capture genetic effects on these cells, which are increasingly being accepted as important in the pathobiology of JIA(3, 41, 42). Neutrophils are not included in the DICE data. At the same time, GTEx provide expression data only at the tissue level, with the exception of a few cell types (e.g., EBV-transformed lymphocytes and cultured skin fibroblasts). Thus, confidently identifying the impact of a given SNP on a particular cell type in the GTEx data set is impossible.

In conclusion, have found that the majority of SNPs identified in an MPRA performed in K562 cells demonstrate associations with expression levels of disease-relevant candidate target genes/tissues using human data from the GTEx and DICE databases. Augmenting MPRA analyses by querying these data sets may facilitate the identification of the breadth of genetic impacts driving risk for JIA and assist in prioritizing the candidates that emerge from MPRA analyses. In the final analysis, identifying the actual risk-driving variants will require further computational sorting of the candidate alleles (e.g., assessing the impact of candidate alleles on DNA topology(1) and/or using machine learning approaches to assess the impact of a given allele on any of discrete phases of transcription(43), followed by unequivocal identification of the impacted genes by (e.g., by CRISPRi/a(18). Finally, the field will need to close the “causal loop” by demonstrating that specific alleles have specific transcriptional effects(44). Because of the labor-intensive nature (and expensed of) such experiments, using human data to inform the design of these experiments is likely to be useful. Ultimately, identifying the impacts of specific alleles on specific cells, tissues, and biological processes is an important step toward using genetic data to stratify patients in clinical trials and for developing precision medicine strategies that will use individual patients’ unique genetic vulnerabilities to guide therapy.

## Data Availability

Data from the original MPRA are available in the Sequence Read Archive (SRA) accession number PRJNA818294.

https://www.ncbi.nlm.nih.gov/sra/?term=PRJNA818294

## Declaration of Generative AI Use

No AI tools were used to create this manuscript or figures.

## Acknowledgements

The authors express their thanks to Tao Liu for computational assistance with the original MPRA.

## Author contributions

KJ – Performed the original MPRA and assisted with the data interpretation and assisted with the preparation of this manuscript.

SK – Assisted in the performance of the original MPRA.

RT – Provided assistance with the design and performance of the original MPRA.

JNJ – Designed the current study and performed the analyses from the DICE and GTEx databases.

